# Effectiveness of BNT162b2 booster doses in England: an observational study in OpenSAFELY-TPP

**DOI:** 10.1101/2022.06.06.22276026

**Authors:** William J Hulme, Elizabeth J Williamson, Elsie Horne, Amelia Green, Linda Nab, Ruth Keogh, Edward PK Parker, Venexia Walker, Tom Palmer, Helen Curtis, Milan Wiedemann, Christine Cunningham, Alex J Walker, Louis Fisher, Brian MacKenna, Christopher T Rentsch, Anna Schultze, Krishnan Bhaskaran, John Tazare, Laurie Tomlinson, Helen I McDonald, Caroline E Morton, Richard Croker, Colm Andrews, Robin Parks, Lisa Hopcroft, Jon Massey, Jessica Morley, Amir Mehrkar, Seb Bacon, Dave Evans, Peter Inglesby, George Hickman, Simon Davy, Iain Dillingham, Tom Ward, Viyasaan Mahalingasivam, Bang Zheng, Ian J Douglas, Stephen JW Evans, Chris Bates, Jonathan AC Sterne, Miguel A Hernán, Ben Goldacre

**Author notes:** equal senior author contribution (Jonathan AC Sterne, Miguel A Hernán, Ben Goldacre).

## Abstract

**Background:** The UK COVID-19 vaccination programme delivered its first “booster” doses in September 2021, initially in groups at high risk of severe disease then across the adult population. The BNT162b2 Pfizer-BioNTech vaccine was used initially, with Moderna mRNA-1273 subsequently also used.

**Methods:** We used the OpenSAFELY-TPP database, covering 40% of English primary care practices and linked to national coronavirus surveillance, hospital episodes, and death registry data, to estimate the effectiveness of boosting with BNT162b2 compared with no boosting in eligible adults who had received two primary course vaccine doses between 16 September and 16 December 2021 when the Delta variant of SARS-CoV-2 was dominant. Follow up was for up to 10 weeks. Each booster recipient was matched with an unboosted control on factors relating to booster priority status and prior immunisation. Additional factors were adjusted for in Cox models estimating hazard ratios (HRs). Outcomes were positive SARS-CoV-2 test, COVID-19 hospitalisation, COVID-19 death and non-COVID-9 death. Booster vaccine effectiveness was defined as 1−HR.

**Results:** Among 4,352,417 BNT162b2 booster recipients matched with unboosted controls, estimated effectiveness of a booster dose compared with two doses only was 50.7% (95% CI 50.1-51.3) for positive SARS-CoV-2 test, 80.1% (78.3-81.8) for COVID-19 hospitalisation, 88.5% (85.0-91.1) for COVID-19 death, and 80.3% (79.0-81.5) for non-COVID-19 death.

Estimated effectiveness was similar among those who had received a BNT162b2 or ChAdOx1-S two-dose primary vaccination course, but effectiveness against severe COVID-19 was slightly lower in those classified as clinically extremely vulnerable (76.3% (73.1-79.1) for COVID-19 hospitalisation, and 85.1% (79.6-89.1) for COVID-19 death). Estimated effectiveness against each outcome was lower in those aged 18-65 years than in those aged 65 and over.

**Conclusion:** Our findings are consistent with strong protection of BNT162b2 boosting against positive SARS-CoV-2 test, COVID-19 hospitalisation, and COVID-19 death.

## Introduction

In mid-September 2021 the national COVID-19 vaccination programme in England administered its first booster doses in adults who had already received their two-dose primary vaccination course (1). Eligibility was initially restricted to those at highest risk of severe disease, then progressively extended. By 15 December 2021 every adult was eligible (2). Booster doses were initially available no earlier than six months after dose two, but this was reduced to three months on 8 December 2021, following concerns over the emergence of the Omicron variant (3) (4). Vaccine prioritisation schedules were guided by recommendations from the Joint Committee for Vaccine and Immunisation (JCVI) expert working group (5).

We aimed to emulate a target trial assessing the effectiveness of booster vaccination with BNT162b2 against COVID-19 outcomes (6) (7), by comparing fully vaccinated adults who did and did not receive a booster vaccine dose. We used the OpenSAFELY-TPP linked primary care database covering around 40% of English residents. The Delta SARS-CoV-2 variant was dominant during the study period.

## Methods

### Data source

All data were linked, stored and analysed securely within the OpenSAFELY platform: https://opensafely.org/. With the approval of NHS England, primary care records managed by the GP software provider TPP were linked, using NHS numbers, to A&E attendance and in-patient hospital spell records via NHS Digital’s Hospital Episode Statistics (HES), national coronavirus testing records via the Second Generation Surveillance System (SGSS), and national death registry records from the Office for National Statistics (ONS). COVID-19 vaccination history is available in the GP record directly via the National Immunisation Management System (NIMS).

### Eligibility criteria

We included adults aged ≥18 years who were eligible for booster vaccination between 16 September and 16 December 2021 inclusive (the study entry period). On each day within this period, eligibility for the study was determined as follows: registered at a GP practice using TPP’s SystmOne clinical information system; received a two-dose primary vaccination course of either BNT162b2 or ChAdOx1-S (mixed dosing and Moderna mRNA-1273 were not considered due to small numbers); not a health or social care worker; not resident in a care or nursing home; not medically housebound or receiving end-of-life care; no evidence of SARS-CoV-2 infection or COVID-19 disease within the previous 90 days; not undergoing an unplanned hospital admission; complete information on sex, ethnicity, deprivation, and NHS region (East of England, Midlands, London, North East and Yorkshire, North West, South East, South West).

Additionally, on days when the rolling weekly average count of BNT162b2 booster doses within strata defined by region, JCVI priority group, and the week of second dose was below 50, study entry did not occur within the stratum. This helped to ensure that study entry was restricted to those eligible and able to receive a booster dose in line with national prioritisation schedules.

### Matching, treatment groups, and follow up

On each day of the study entry period, each eligible person receiving a booster dose was recruited to the treatment group and matched, if possible, with an eligible unboosted control person. Pairs were matched on: primary course vaccine brand; date of second vaccine dose, within 7 days; NHS region; evidence of prior SARS-CoV-2 infection (positive SARS-CoV-2 test, “probable” infection documented in primary care, or COVID-19 hospitalisation); clinical risk groups used for prioritisation (clinically extremely vulnerable, clinically at-risk, neither); age within 3 years; and age groups used by JCVI for prioritisation. People selected as controls were not eligible to be included again as a control, but were eligible for subsequent inclusion in the booster group. Any unmatched boosted people were excluded.

Each person was followed from the day of study entry (i.e., time zero) until the earliest of outcome, death, practice de-registration, 10 weeks, or 31 December 2021. Additionally, follow-up was censored if the matched control was boosted, for both boosted and unboosted groups.

### Outcomes

Four outcomes were considered: positive SARS-CoV-2 test; COVID-19 hospitalisation; COVID-19 death; and non-COVID-19 death. SARS-CoV-2 infections were identified using SGSS testing records and based on swab date. Both polymerase chain reaction (PCR) and lateral flow test results are included, without differentiation between symptomatic and asymptomatic infection. COVID-19 hospitalisations were identified using HES in-patient hospital records with U07.1 or U07.2 reason for admission ICD-10 codes. Deaths were classified as from COVID-19 if deaths with the same ICD-10 codes were mentioned anywhere on the death certificate (i.e., as an underlying or contributing cause of death).

## Statistical Analysis

We estimated Kaplan-Meier cumulative incidence curves separately in the boosted and unboosted groups, and estimated the 10-week risk differences based on these. We used Cox models, stratified by study entry day, NHS region, and two-dose primary vaccine course, to estimate hazard ratios (HRs) comparing boosted with unboosted people, overall and separately for days 1-28 and 29-70. We also estimated HRs within shorter time periods (days 1-7, 8-14, 15-28, 29-42, and 43-70).

As we were not able to match on all potential confounders, these models included the following additional covariates: sex (male or female); English Index of Multiple Deprivation (IMD, grouped by quintile); ethnicity (Black, Mixed, South Asian, White, Other, as per the UK census); prior morbidity count (diabetes, BMI over 40kg/m^2^, chronic heart disease, chronic kidney disease, chronic liver disease, chronic respiratory disease or severe asthma, chronic neurological disease); learning disabilities; severe mental illness; the number of SARS-CoV-2 tests in the 6 months prior to the study start date; elective in-hospital episode at baseline. There were no missing values in these covariates, because they were defined by the presence or absence of clinical codes or events. A robust variance estimator was used to account for inclusion of some people in both the boosted and unboosted groups.

Booster vaccine effectiveness was calculated as 1-HR, expressed as a percentage. We also estimated booster effectiveness separately in the following subgroups: primary vaccine course; age (18-64 or ≥65 years); clinically extremely vulnerable or not; evidence of prior SARS-CoV-2 infection or not.

### Software, code, and reproducibility

Data management and analyses were conducted in Python version 3.8.10 and R version 4.0.2. All code is shared openly for review and re-use under MIT open license at https://github.com/opensafely/booster-effectiveness. The supplementary materials provide further details of the codelists and data sources used for all variables in the study. Detailed pseudonymised patient data is potentially re-identifiable and therefore not shared. Any reported figures based on counts below 6 are redacted or rounded for disclosure control. This study followed the STROBE-RECORD reporting guidelines.

### Patient and public involvement

We have developed a publicly available website https://opensafely.org/ through which we invite any patient or member of the public to contact us regarding this study or the broader OpenSAFELY project.

## Results

### Study population and matching

7,339,110 adults registered at a TPP practice received a BNT162b2 booster vaccination during the study entry period, with 5,095,279 (69.4%) eligible for inclusion in the treatment group, of whom 4,352,417 (85.4%) were matched with unboosted controls (Figure 1). 1,714,615 people were matched as controls then rematched in the booster group, resulting in 6,990,219 people included in the study in total.

**Figure 1:**
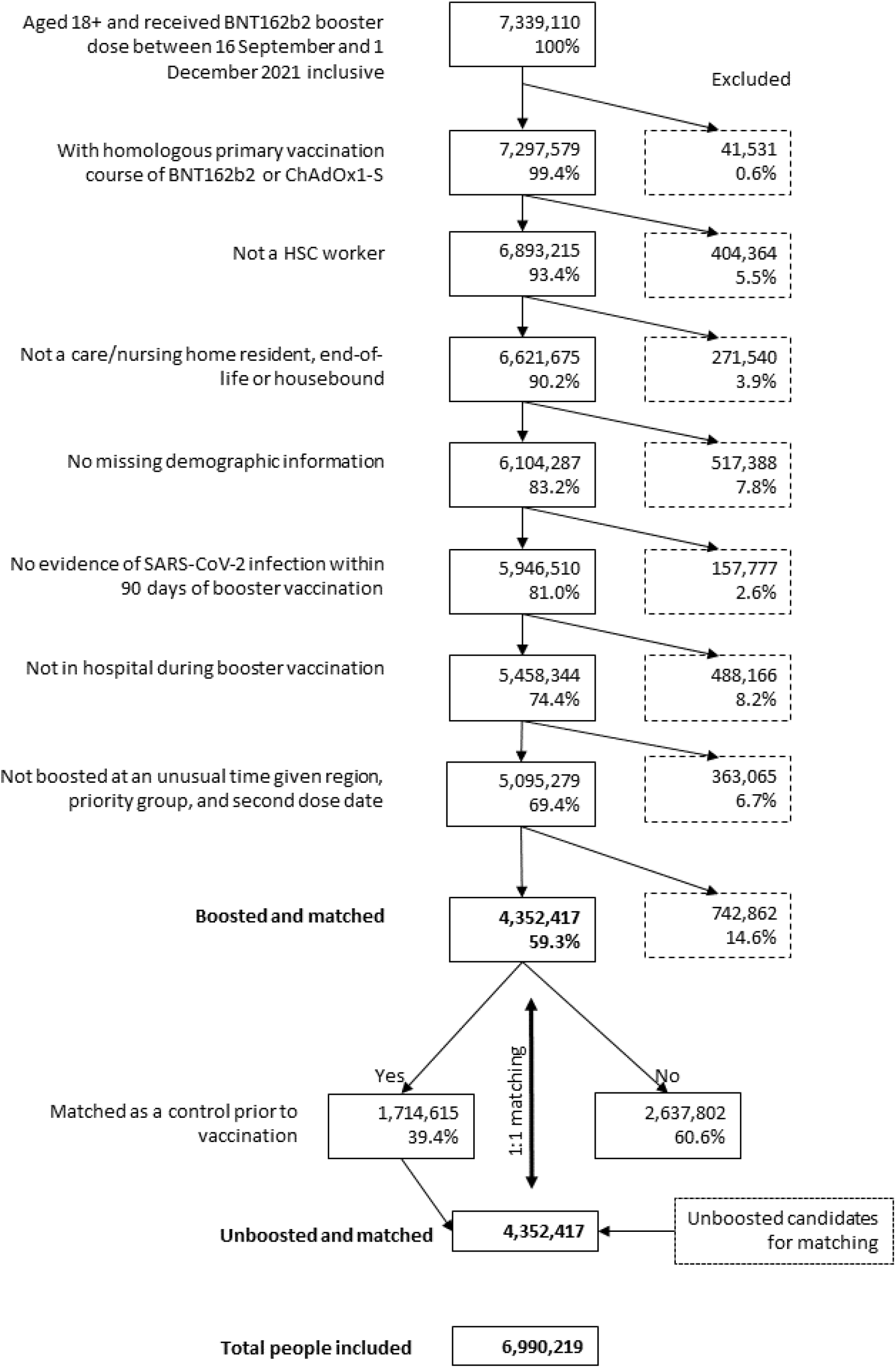
Inclusion criteria

As expected, the matching factors were identically distributed in the boosted and unboosted groups at the start of follow up (Table 1), and the proportion of people with prior morbidities was generally similar between the groups. However, unboosted controls had higher levels of deprivation, higher rates of learning disabilities and severe mental illness, and lower SARS-CoV-2 testing frequency, though the standardised mean difference was consistently below 0.1 (Supplementary Figure S2).

**Table 1:**
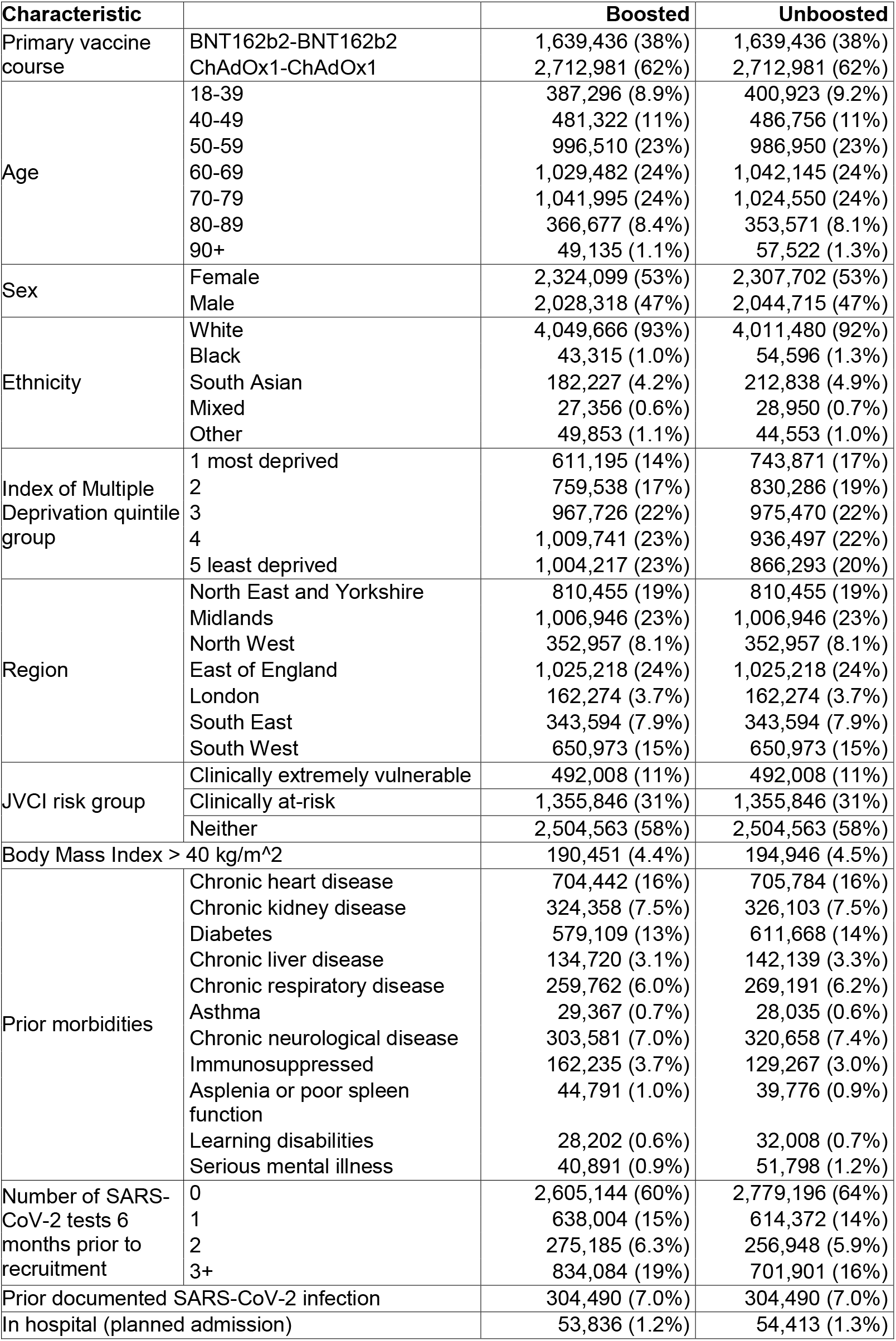
Characteristics (%) of 4,352,417 boosted and 4,352,417 matched unboosted participants on the day of study entry.

**Table 2:**
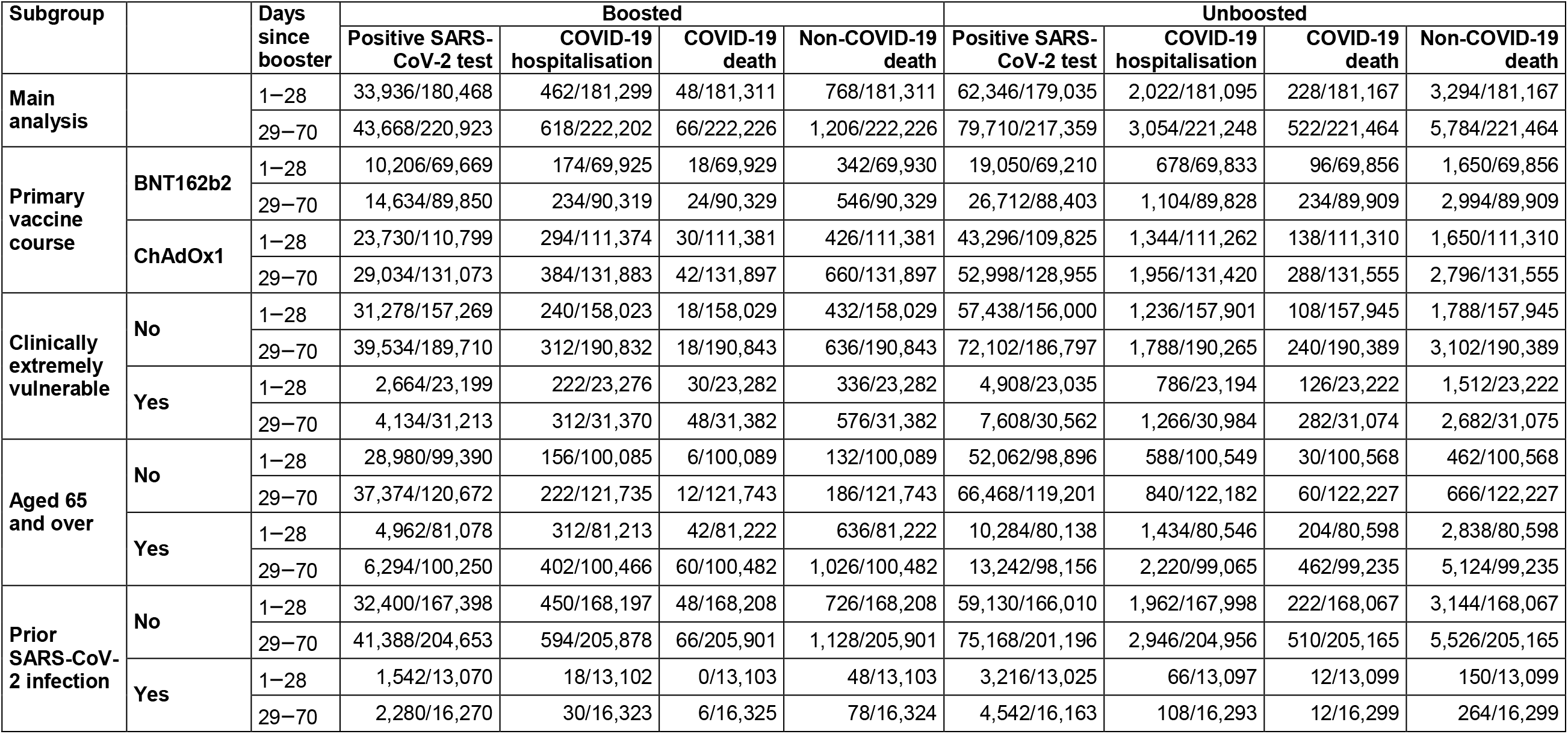
Total events and person-years of follow-up, for each outcome, in the boosted and unboosted groups.

### Estimated booster effectiveness

#### Main analysis

There were 123,378 positive SARS-CoV-2 tests, 3,672 COVID-19 hospitalisations, 588 COVID-19 deaths, and 6,990 non-COVID-19 deaths across 23,151,145 person-weeks of follow-up. The rate of increase in the cumulative incidence of positive SARS-CoV-2 tests in boosted people tapered substantially around 7 days after boosting, something not observed in unboosted controls (Figure 2). Similar patterns were not observed for more severe outcomes, and there were differences in the cumulative incidence between the groups apparent in the first few days.

**Figure 2:**
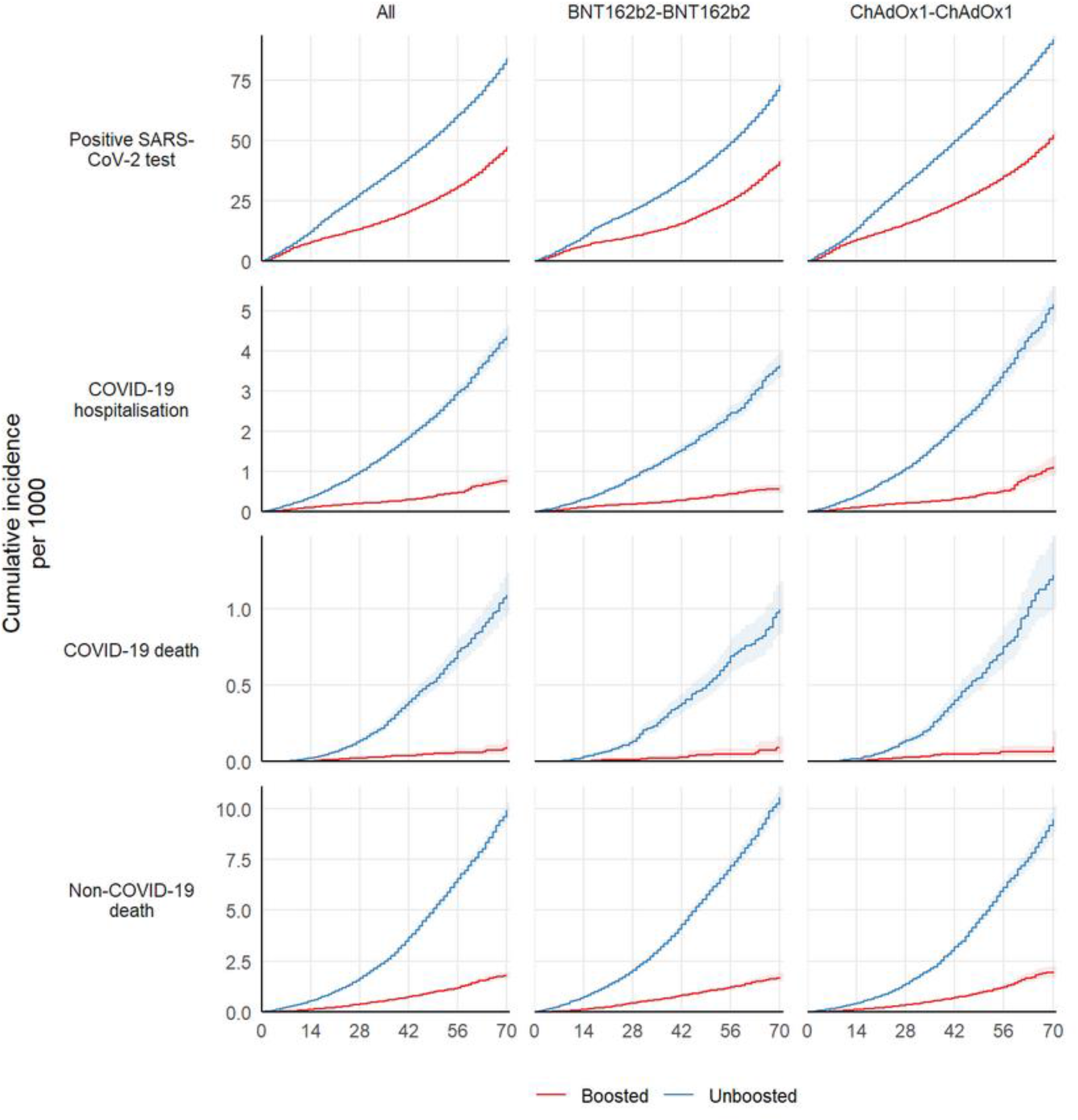
Kaplan-Meier estimates of cumulative incidence in matched boosted and unboosted treatment groups, stratified by primary course and without further adjustment for potential confounders.

The 10-week risk of positive SARS-CoV-2 test was 47.3 and 84.0 per 1000 in the boosted group and unboosted groups respectively (risk difference -36.8 per 1000). Corresponding 10-week risks per 1000 were 0.8 and 4.4 for COVID-19 hospitalisation (risk difference - 3.6), 0.1 and 1.1 for COVID-19 death (-1.0), and 1.8 and 9.9 for non-COVID-19 death (-8.1).

The overall (days 1-70) estimated booster effectiveness comparing the boosted and unboosted groups was 50.7% (95% CI 50.1-51.3) for positive SARS-CoV-2 test, 80.1% (78.3-81.8) for COVID-19 hospitalisation, 88.5% (85.0-91.1) for COVID-19 death, and 80.3% (79.0-81.5) for non-COVID-19 death (Table 3). Estimated booster effectiveness against COVID-19 outcomes was generally lower during days 1-28 than days 29-70: 49.3% (48.6-50.0) and 52.7% (51.5-53.8) for respectively positive SARS-CoV-2 test; 76.8% (74.3-79.0) and 85.1% (82.4-87.4) respectively for COVID-19 hospitalisation; 81.2% (74.0-86.5) and 93.4% (89.8-95.8) respectively for COVID-19 death; and 77.6% (75.8-79.3) and 83.5% (81.7-85.1) for non-COVID-19 death.

**Table 3:** Estimated booster vaccine effectiveness (100 × (1 − hazard ratio)) for the main and subgroup analyses and for all outcomes.

| Outcome | Subgroup |  | Booster vaccine effectiveness (95% CI) |  |  |
| --- | --- | --- | --- | --- | --- |
|  |  |  | 1 - 28 days | 29 - 70 days | 1 - 70 days |
| Positive SARS-CoV-2 test | All |  | 49.3 (48.6-50.0) | 52.7 (51.5-53.8) | 50.7 (50.1-51.3) |
|  | Primary course vaccine brand | BNT162b2 | 49.6 (48.3-50.8) | 49.7 (47.8-51.5) | 50.2 (49.1-51.2) |
|  |  | ChAdOx1 | 49.2 (48.4-50.0) | 54.8 (53.3-56.3) | 51.0 (50.2-51.7) |
|  | Clinically Extremely Vulnerable | No | 49.4 (48.7-50.1) | 52.5 (51.2-53.7) | 50.7 (50.0-51.3) |
|  |  | Yes | 48.1 (45.6-50.5) | 53.2 (50.1-56.1) | 50.8 (48.8-52.6) |
|  | Age (years) | 18-64 | 48.4 (47.6-49.1) | 50.9 (49.5-52.2) | 49.5 (48.8-50.1) |
|  |  | 65 and over | 51.4 (49.7-53.1) | 59.1 (56.3-61.7) | 55.1 (53.7-56.5) |
|  | Prior SARS-CoV-2 infection | No | 49.3 (48.6-50.0) | 53.2 (51.9-54.4) | 50.8 (50.2-51.4) |
|  |  | Yes | 54.1 (51.2-56.8) | 49.4 (44.6-53.7) | 53.4 (51.0-55.7) |
| COVID-19 hospitalisation | All |  | 76.8 (74.3-79.0) | 85.1 (82.4-87.4) | 80.1 (78.3-81.8) |
|  | Primary course vaccine brand | BNT162b2 | 73.8 (68.9-77.9) | 86.1 (81.9-89.4) | 79.3 (76.1-82.1) |
|  |  | ChAdOx1 | 78.2 (75.2-80.8) | 83.9 (80.0-87.1) | 80.6 (78.3-82.6) |
|  | Clinically Extremely Vulnerable | No | 79.9 (76.8-82.5) | 87.6 (84.1-90.4) | 82.9 (80.7-84.9) |
|  |  | Yes | 71.7 (67.1-75.6) | 82.5 (78.0-86.1) | 76.3 (73.1-79.1) |
|  | Age (years) | 18-64 | 74.2 (69.2-78.4) | 73.3 (64.6-79.8) | 74.8 (70.7-78.3) |
|  |  | 65 and over | 77.8 (74.9-80.4) | 88.5 (85.7-90.7) | 82.1 (80.1-83.9) |
|  | Prior SARS-CoV-2 infection | No | 76.8 (74.3-79.1) | 85.1 (82.2-87.5) | 80.2 (78.4-81.9) |
|  |  | Yes | 72.8 (52.4-84.5) | 80.0 (58.1-90.4) | 78.4 (66.4-86.1) |
| COVID-19 death | All |  | 81.2 (74.0-86.5) | 93.4 (89.8-95.8) | 88.5 (85.0-91.1) |
|  | Primary course vaccine brand | BNT162b2 | 100 | 92.6 (86.5-96.0) | 90.5 (85.6-93.7) |
|  |  | ChAdOx1 | 77.4 (66.3-84.9) | 93.5 (87.6-96.6) | 86.6 (81.3-90.4) |
|  | Clinically Extremely Vulnerable | No | 100 | 100 | 92.8 (88.4-95.6) |
|  |  | Yes | 77.4 (66.1-84.9) | 89.8 (83.1-93.8) | 85.1 (79.6-89.1) |
|  | Age (years) | 18-64 | 100 | 93.7 (73.8-98.5) | 86.3 (71.4-93.4) |
|  |  | 65 and over | 82.0 (74.4-87.3) | 93.2 (89.2-95.7) | 88.5 (84.8-91.3) |
|  | Prior SARS-CoV-2 infection | No | 80.6 (73.1-86.1) | 93.9 (90.4-96.2) | 88.6 (85.1-91.2) |
|  |  | Yes | 100 | -Inf | 90.0 (29.3-98.6) |
| Non-COVID-19 death | All |  | 77.6 (75.8-79.3) | 83.5 (81.7-85.1) | 80.3 (79.0-81.5) |
|  | Primary course vaccine brand | BNT162b2 | 79.8 (77.3-82.0) | 85.4 (83.1-87.4) | 82.5 (80.8-84.0) |
|  |  | ChAdOx1 | 75.2 (72.4-77.7) | 81.2 (78.3-83.7) | 77.8 (75.8-79.6) |
|  | Clinically Extremely Vulnerable | No | 76.6 (74.0-78.9) | 85.7 (83.4-87.6) | 80.6 (78.9-82.2) |
|  |  | Yes | 78.5 (75.7-80.9) | 81.4 (78.6-83.8) | 80.0 (78.1-81.8) |
|  | Age (years) | 18-64 | 72.7 (66.8-77.6) | 78.9 (71.1-84.6) | 75.0 (70.4-78.9) |
|  |  | 65 and over | 78.2 (76.2-80.0) | 83.3 (81.4-85.0) | 80.5 (79.1-81.8) |
|  | Prior SARS-CoV-2 infection | No | 77.9 (76.0-79.6) | 84.0 (82.2-85.6) | 80.6 (79.3-81.8) |
|  |  | Yes | 72.9 (62.0-80.7) | 73.7 (61.6-81.9) | 73.4 (65.7-79.4) |

### Subgroup analyses

Outcomes were broadly similar between subgroups defined by primary vaccine course. For those who had received BNT162b2, estimated booster effectiveness was 50.2% (49.1-51.2) for positive SARS-CoV-2 test, 79.3% (76.1-82.1) for COVID-19 hospitalisation, 90.5% (85.6-93.7) for COVID-19 death, and 82.5% (80.8-84.0) for non-COVID-19 death. Corresponding estimates for ChAdOx1-S recipients were 51.0% (50.2-51.7), 80.6% (78.3-82.6), 86.6% (81.3-90.4), and 77.8% (75.8-79.6) respectively.

Among people classified as clinically extremely vulnerable, estimated booster effectiveness was 50.8% (48.8-52.6) for positive SARS-CoV-2 test, 76.3% (73.1-79.1) for COVID-19 hospitalisation, and 85.1% (79.6-89.1) for COVID-19 death. Among people not classified as clinically extremely vulnerable estimated booster effectiveness was slightly higher, at 50.7% (50.0-51.3), 82.9% (80.7-84.9) and 92.8% (88.4-95.6) respectively. Estimated booster effectiveness for non-COVID-19 death was similar for those who were and were not classified as clinically extremely vulnerable: 80.0% (78.1-81.8) and 80.6% (78.9-82.2) respectively.

Among people aged ≥65 years, estimated booster effectiveness was 55.1% (53.7-56.5) for positive SARS-CoV-2 test, 82.1% (80.1-83.9) for COVID-19 hospitalisation, 88.5% (84.8-91.3) for COVID-19 death, and 80.5% (79.1-81.8) for non-COVID-19 death. Corresponding effectiveness appeared lower in people aged 18-64 years: 49.5% (48.8-50.1) for positive SARS-CoV-2 tests, 74.8% (70.7-78.3) for COVID-19 hospitalisation, 86.3% (71.4-93.4) for COVID-19 death, and 75.0% (70.4-78.9) for non-COVID-19 death.

Outcomes were similar for COVID-19-related events when comparing people with or without evidence of prior SARS-CoV-2 infection. Estimated booster effectiveness for those with evidence of prior infection was 53.4% (51.0-55.7) for positive SARS-CoV-2 test, 78.4% (66.4-86.1) for COVID-19 hospitalisation, 90.0% (29.3-98.6) for COVID-19 death. Corresponding estimates in people with no evidence of prior infection were 50.8% (50.2-51.4), 80.2% (78.4-81.9), and 88.6% (85.1-91.2), respectively. For non-COVID-19 death, estimated booster effectiveness was slightly lower in those with prior evidence of SARS-CoV-2 infection (73.4% (65.7-79.4)) than in those without (80.6% (79.3-81.8)).

When booster effectiveness was estimated within shorter time periods, effectiveness against positive SARS-CoV-2 tests appeared to wane towards the end of follow up (68.2% (95% CI 67.4-69.0) during days 15-28 versus 45.3% (43.3-47.1) during days 43-70). This pattern was observed in each subgroup defined by primary course vaccine brand (Figure 3). For more severe outcomes, period-specific effectiveness estimates were imprecisely estimated but broadly similar between primary course vaccine brands.

**Figure 3:**
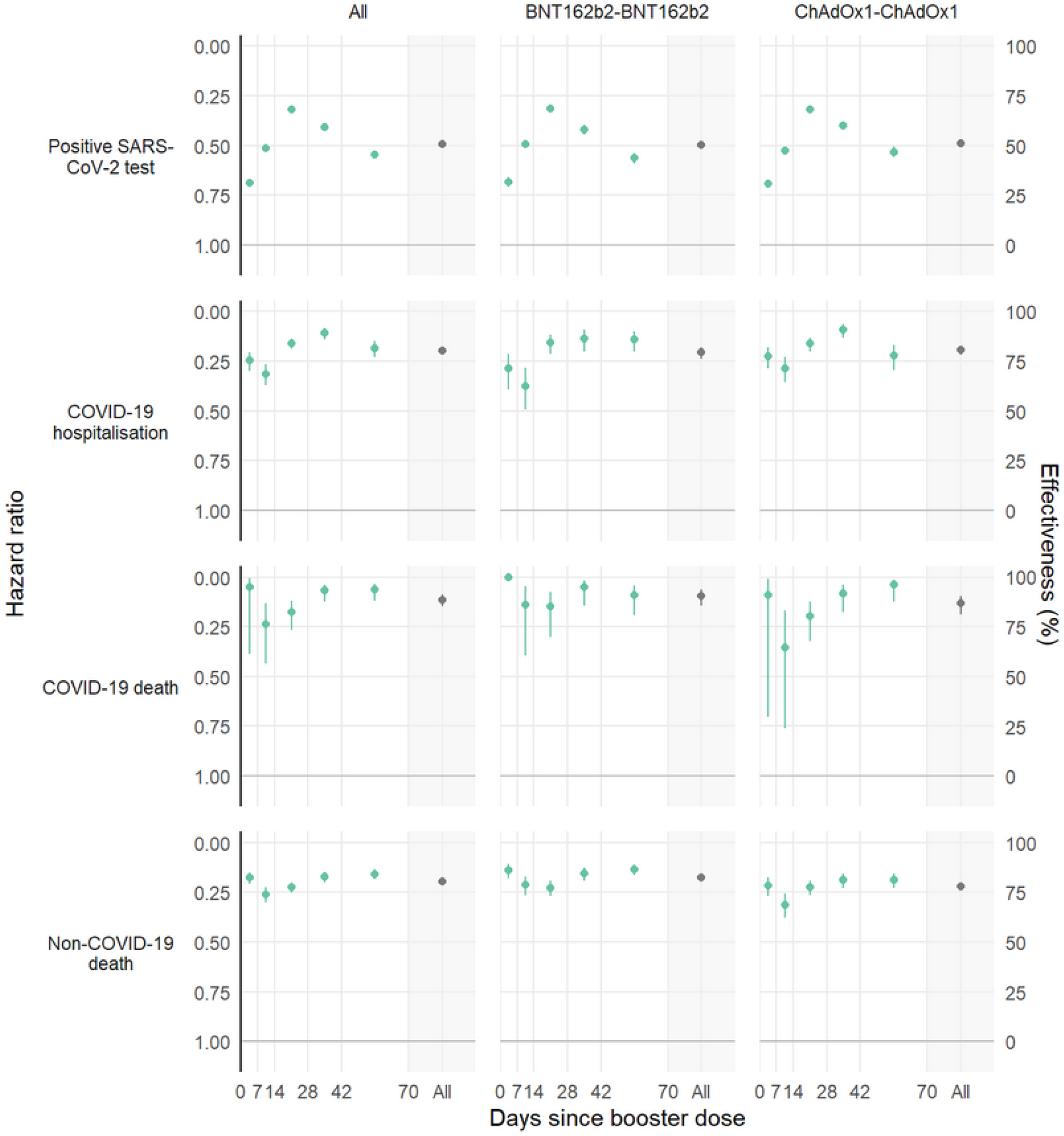
Estimated booster vaccine effectiveness (100 × (1 − hazard ratio)) for each outcome based on the fully adjusted model, stratified by primary course and time since boosting. Models with less extensive confounder adjustment are provided in supplementary materials (Figure S3).

## Discussion

In this observational cohort study based on nearly 7 million adults in England during a period when the Delta variant was dominant (8), we estimated that a BNT162b2 booster dose reduced rates of positive SARS-CoV-2 tests by approximately 50% over the first 10 weeks after booster vaccination. We estimated substantial reductions in rates of COVID-19 hospitalisation, COVID-19 death, and non-COVID-19 death. Estimated booster effectiveness against these severe outcomes appeared similar regardless of whether the primary vaccine course was with ChAdOx1-S or BNT162b2, and whether there was evidence of prior infection or not. However, the estimated protection against severe COVID-19 was lower in people classified as clinically extremely vulnerable compared with those not so classified, and lower in those aged under 65 years. Protection against positive SARS-CoV-2 tests appeared to wane from around 6 weeks after booster vaccination.

### Strengths and Limitations

Like in any observational study, our estimates could be confounded if the boosted and the unboosted groups had different risks at the time of booster vaccination. However, our use of routinely-collected electronic health care records, with comprehensive coverage of primary care and hospitalisations, enabled us to carefully match booster recipients with unboosted controls on multiple characteristics and to further adjust for additional potential confounding factors.

Due to the observational nature of the study, people with undiagnosed SARS-CoV-2 infection or COVID-19 symptoms, which are not routinely recorded, could not be comprehensively excluded at baseline. These people are more likely to be included in the unboosted group, as people with symptoms were told to defer booster vaccination. This may explain the premature effectiveness that we estimated against hospitalisation and COVID-19 death during the first week. However, this bias is transient by definition and should not affect our long-term estimates.

We could not fully adjust for smoking and other lifestyle factors that are associated with increased all-cause mortality. This might explain at least part of the strong protection of booster vaccination that we estimated against non-COVID-19 death. However, it is unclear whether these general risk factors would also confound the estimates of effectiveness against COVID-19 related outcomes. COVID-19 vaccination, including booster vaccination, may protect against deaths that are not coded as due to COVID-19 by preventing harms of severe COVID-19, such as acute myocardial infarction and stroke(9,10). We are unable to compare our non-COVID-19 death results with other recent studies of booster effectiveness, because those studies did not report on non-COVD-19 death.

Positive SARS-CoV-2 test data likely underestimates the true incidence of infection. Both lateral flow tests and PCR tests were freely available in the UK during the study period, but many asymptomatic infections and some symptomatic infections will not have been recorded in national data, despite encouragement to report results of lateral flow tests and seek a confirmatory PCR test when these were positive. Potential differences in testing behaviour between boosted and unboosted people may also undermine the reliability of testing data as a means to assess effectiveness (11). SARS-CoV-2 testing was not widely available early in the pandemic so prior infection is also likely to be under-ascertained. Hospital admission records are only completed after discharge, so some very long, and infrequent, hospital stays commencing within the follow-up period may not have been included.

We excluded a number of groups, such as health care workers and care home residents, where testing use, vaccination uptake, and infection risk were unusual or had substantial within-group heterogeneity that could not adequately measured and controlled for. The generalisability of our results to these excluded groups is unclear. Due to small numbers, we did not study booster effectiveness in those who had received the mRNA-1273 vaccine as their primary course or those who had received a heterologous primary course.

### Findings in context

A phase III trial in two-dose BNT162b2 recipients without prior infection reported a relative boster vaccination efficacy over a median of 2.5 months of 95.6% (95% CI 89.3-98.6) against Delta infection (12). Observational studies in the UK have used ‘test-negative’ designs to provide evidence of the effectiveness of booster vaccination against symptomatic COVID-19 infection, in comparison to second dose recipients. Andrews et al. reported estimates of relative vaccine effectiveness in the first 14 days after BNT162b2 booster vaccination of 87.4% (95%CI 84.9-89.4) where the primary course was ChAdOx1-S and 84.4% (95%CI 82.8-85.8) for BNT162b2, in those aged over 50 years (13). A study in Scotland reported comparable estimates of relative vaccine effectiveness for S gene positive (a surrogate the Delta variant in the latter half of 2021 in the United Kingdom) symptomatic infection (83% (95%CI 81-84) 6-49 years, 88% (95%CI 86-89) over 50 years) in the 14 days after a BNT162b2 or mRNA-1273 booster dose, but lower estimates for S gene negative symptomatic infection (56% (95%CI 51-60) 6-49 years, 57% (95%CI 52-62) over 50 years) (14). We assessed effectiveness of booster vaccination against test positivity, combining both asymptomatic and symptomatic testing from documented PCR and lateral flow tests, and estimated substantially lower protection for this outcome compared with symptomatic infection only as previously estimated using test-negative designs in UK data. This suggests that booster vaccination is effective at reducing disease severity but not solely by reducing susceptibility to infection itself. It may also suggest vulnerability of the test-negative design to bias when booster vaccination reduces disease severity (15).

A study in Israeli health registry data using 1:1 matching found strong protection against admission (1–risk ratio (95%CI) = 93% (88-97)), severe disease (92% (82-97)), and COVID-19 death (81% (59-97)), with similar results in specific demographic and clinical subgroups (7). Our study, which also used matching but additionally adjusted for potential confounders not included as matching factors, found slightly lower estimates of effectiveness against hospitalization but higher estimates for death.

Our results also provide important additional insights into effectiveness in clinical subgroups, younger age groups, and more severe outcomes over a longer period of follow up. Encouragingly, we found effectiveness was broadly similar irrespective of whether BNT162b2 or ChAdOx1-S was given for the primary vaccination course, and there were only slight reductions in effectiveness in those who were clinically extremely vulnerable or aged under 65 years. The absence of large discrepancies in booster effectiveness in different subgroups may simplify the coordination of future booster campaigns in the UK, allowing boosting to be prioritised on vulnerability to infection and severe disease without consideration for differential vaccine escape.

## Conclusion

This study of almost 7 million people in England estimated high protection for BNT162b2 boosting against positive SARS-CoV-2 tests, COVID-19 hospitalisation, and death during a period in which the Delta variant was dominant. Some evidence of waning effectiveness against positive SARS-CoV-2 tests suggests the need for careful monitoring of booster vaccine effectiveness over time.

## Data Availability

Detailed pseudonymised patient data is potentially re-identifiable and therefore not shared.

https://github.com/opensafely/booster-effectiveness/tree/713c93530701a1495b2cc742a1c6a1c24419880d

## Acknowledgements

We are very grateful for all the support received from the TPP Technical Operations team throughout this work, and for generous assistance from the information governance and database teams at NHS England / NHSX.

Our thanks to Susana Monge-Corella for helpful discussions on optimising the matching algorithm.

## Funding

This work was jointly funded by UKRI [COV0076;MR/V015737/1], the Longitudinal Health and Wellbeing strand of the National Core Studies programme (MC_PC_20030; MC_PC_20059; COV-LT-0009), NIHR and Asthma UK-BLF. The OpenSAFELY data science platform is funded by the Wellcome Trust (222097/Z/20/Z).

BG’s work on better use of data in healthcare more broadly is currently funded in part by: the Bennett Foundation, the Wellcome Trust, NIHR Oxford Biomedical Research Centre, NIHR Applied Research Collaboration Oxford and Thames Valley, the Mohn-Westlake Foundation; all Bennett Institute staff are supported by BG’s grants on this work. AS is employed by LSHTM on a fellowship sponsored by GSK. KB holds a Wellcome Senior Research Fellowship (220283/Z/20/Z). HIM is funded by the National Institute for Health Research (NIHR) Health Protection Research Unit in Immunisation, a partnership between UK Health Security Agency and LSHTM. BMK is also employed by NHS England working on medicines policy and clinical lead for primary care medicines data. EW holds grants from MRC. ID holds grants from NIHR and GSK.

The views expressed are those of the authors and not necessarily those of the NIHR, NHS England, UK Health Security Agency (UKHSA) or the Department of Health and Social Care.

Funders had no role in the study design, collection, analysis, and interpretation of data; in the writing of the report; and in the decision to submit the article for publication.

For the purpose of Open Access, the author has applied a CC BY public copyright licence to any Author Accepted Manuscript (AAM) version arising from this submission.

### Information governance and ethical approval

NHS England is the data controller for OpenSAFELY-TPP; TPP is the data processor; and study authors using OpenSAFELY have the approval of NHS England. This implementation of OpenSAFELY is hosted within the TPP environment which is accredited to the ISO 27001 information security standard and is NHS IG Toolkit compliant (16) (17) ; Patient data has been pseudonymised for analysis and linkage using industry standard cryptographic hashing techniques; all pseudonymised datasets transmitted for linkage onto OpenSAFELY are encrypted; access to the platform is via a virtual private network (VPN) connection, restricted to a small group of researchers; the researchers hold contracts with NHS England and only access the platform to initiate database queries and statistical models; all database activity is logged; only aggregate statistical outputs leave the platform environment following best practice for anonymisation of results such as statistical disclosure control for low cell counts (18). The OpenSAFELY research platform adheres to the obligations of the UK General Data Protection Regulation (GDPR) and the Data Protection Act 2018. In March 2020, the Secretary of State for Health and Social Care used powers under the UK Health Service (Control of Patient Information) Regulations 2002 (COPI) to require organisations to process confidential patient information for the purposes of protecting public health, providing healthcare services to the public and monitoring and managing the COVID-19 outbreak and incidents of exposure; this sets aside the requirement for patient consent (19). Taken together, these provide the legal bases to link patient datasets on the OpenSAFELY platform. General practices, from which the primary care data are obtained, are required to share relevant health information to support the public health response to the pandemic, and have been informed of the OpenSAFELY analytics platform.

This study was approved by the Health Research Authority (REC reference 20/LO/0651) and by the LSHTM Ethics Board (reference 21863).

JACS and BG are guarantors.

## References

1. NHS England NHS begins COVID-19 booster vaccination campaign. https://www.england.nhs.uk/2021/09/nhs-begins-covid-19-booster-vaccination-campaign/;

2. NHS England NHS booster bookings open to every eligible adult. https://www.england.nhs.uk/2021/12/nhs-booster-bookings-open-to-every-eligible-adult/;

3. NHS England NHS people 40 and over to get their lifesaving booster jab three months on from second dose. https://www.england.nhs.uk/2021/12/people-40-and-over-to-get-their-lifesaving-booster-jab-three-months-on-from-second-dose/;

4. NHS England NHS to roll out life-saving booster jab to people aged 30-plus. https://www.england.nhs.uk/2021/12/nhs-to-roll-out-life-saving-booster-jab-to-people-aged-30-plus/;

5. COVID-19: the green book, chapter 14a [Internet]. Available from: https://www.gov.uk/government/publications/covid-19-the-green-book-chapter-14a

6. Dagan N, Barda N, Kepten E, Miron O, Perchik S, Katz MA, et al. BNT162b2 mRNA Covid-19 Vaccine in a Nationwide Mass Vaccination Setting. New England Journal of Medicine [Internet]. 2021 Apr 15;384(15):1412–23. Available from: http://dx.doi.org/10.1056/NEJMoa2101765

7. Barda N, Dagan N, Cohen C, Hernán MA, Lipsitch M, Kohane IS, et al. Effectiveness of a third dose of the BNT162b2 mRNA COVID-19 vaccine for preventing severe outcomes in Israel: an observational study. The Lancet [Internet]. 2021 Dec;398(10316):2093–100. Available from: http://dx.doi.org/10.1016/S0140-6736(21)02249-2

8. COVID-19: Omicron daily overview. GOV.UK. https://www.gov.uk/government/publications/covid-19-omicron-daily-overview;

9. Tan BK, Mainbourg S, Friggeri A, Bertoletti L, Douplat M, Dargaud Y, et al. Arterial and venous thromboembolism in COVID-19: a study-level meta-analysis. Thorax [Internet]. 2021 Feb 23;76(10):970–9. Available from: http://dx.doi.org/10.1136/thoraxjnl-2020-215383

10. Knight R, Walker V, Ip S, Cooper JA, Bolton T, Keene S, et al. Association of COVID-19 with arterial and venous vascular diseases: A population-wide cohort study of 48 million adults in england and wales [Internet]. 2021. Available from: http://dx.doi.org/10.1101/2021.11.22.21266512

11. Glasziou P, McCaffery K, Cvejic E, Batcup C, Ayre J, Pickles K, et al. Testing behaviour may bias observational studies of vaccine effectiveness [Internet]. 2022. Available from: http://dx.doi.org/10.1101/2022.01.17.22269450

12. Pfizer and BioNTech Announce Phase 3 Trial Data Showing High Efficacy of a Booster Dose of Their COVID-19 Vaccine. https://www.businesswire.com/news/home/20211021005491/en/Pfizer-and-BioNTech-Announce-Phase-3-Trial-Data-Showing-High-Efficacy-of-a-Booster-Dose-of-Their-COVID-19-Vaccine; 2021.

13. Andrews N, Stowe J, Kirsebom F, Toffa S, Sachdeva R, Gower C, et al. Effectiveness of COVID-19 booster vaccines against COVID-19-related symptoms, hospitalization and death in England. Nature Medicine [Internet]. 2022 Jan 14;28(4):831–7. Available from: http://dx.doi.org/10.1038/s41591-022-01699-1

14. Sheikh A, Kerr S, Woolhouse M, McMenamin J, Robertson C. Severity of Omicron variant of concern and vaccine effectiveness against symptomatic disease: National cohort with nested test negative design study in Scotland. 2021 Dec;

15. Lewnard JA, Tedijanto C, Cowling BJ, Lipsitch M. Measurement of Vaccine Direct Effects Under the Test-Negative Design. American Journal of Epidemiology [Internet]. 2018 Aug 7;187(12):2686–97. Available from: http://dx.doi.org/10.1093/aje/kwy163

16. BETA Data Security Standards [Internet]. Available from: https://digital.nhs.uk/about-nhs-digital/our-work/nhs-digital-data-and-technology-standards/framework/beta---data-security-standards

17. Data Security and Protection Toolkit [Internet]. Available from: https://digital.nhs.uk/data-and-information/looking-after-information/data-security-and-information-governance/data-security-and-protection-toolkit

18. ISB1523: Anonymisation Standard for Publishing Health and Social Care Data [Internet]. Available from: https://digital.nhs.uk/data-and-information/information-standards/information-standards-and-data-collections-including-extractions/publications-and-notifications/standards-and-collections/isb1523-anonymisation-standard-for-publishing-health-and-social-care-data

19. Coronavirus (COVID-19): Notification to organisations to share information - GOV.UK [Internet]. 2020. Available from: https://web.archive.org/web/20200421171727/ https://www.gov.uk/government/publications/coronavirus-covid-19-notification-of-data-controllers-to-share-information

