## Supplementary material for "Effectiveness of BNT162b2 booster doses in England: an observational study in OpenSAFELY-TPP": supplmentary material: supplement.html

Effectiveness of COVID-19 Booster doses in England: a sequential trials study in OpenSAFELY: Supplementary materials


### Effectiveness of COVID-19 Booster doses in England: a sequential trials study in OpenSAFELY: Supplementary materials

### COVID-19 vaccination history

The dates and brands of any COVID-19 vaccines administered as part of
the national roll-out (e.g., excluding in vaccine trials and vaccines
administered abroad) are available in the primary care records via
transfer from the National Immunisation Management System (NIMS).
Vaccine cardinality (dose 1, dose 2, …) is determined by vaccination
dates alone. We cannot distinguish between “third” doses in
immunosuppressed or otherwise vulnerable people which were administered
ahead of the national booster roll-out, and “booster” doses, i.e.,
indication for 3rd vaccine dose. However, we match on characteristics
that approximate determinants of third dose vaccination, and the
recruitment period excludes a majority of early third dose
recipients.

### Matching details

Booster eligibility was dependent on vaccine priority group, largely
determined by clinical vulnerability and age. Booster doses were
initially available 6 months after administration of the second dose,
later reduced to 3 months following concerns over the surge in cases and
the emergence of the seemingly more transmissible Omicron variant.
Determining vaccine eligibility for a given person on a given day is
therefore complex. To avoid these complexities, we determined trial
eligibility for each person on each day of recruitment by ensuring that
there were sufficiently many people with similar characteristics who
were being vaccinated on that day. Specifically, a rolling weekly
average was calculated within strata defined by region, priority group,
and week of second vaccine dose. If fewer than 50 people had been
vaccinated within each strata on each day, then that strata was not
eligible for recruitment on that day.

The Joint Committee on Vaccine and Immunisation (JCVI) priority
groups were defined as follows:

| Priority group | Description |
| --- | --- |
| 1 | Residents in a care home for older adults Staff working in care homes for older adults |
| 2 | All those 80 years of age and over Frontline health and social care workers |
| 3 | All those 75-79 years of age |
| 4 | All those 70-74 years of age Individuals aged 16-69 in a high risk group |
| 5 | All those 65-69 years of age |
| 6 | Adults aged 16-64 years in an at-risk group |
| 7 | All those 60-64 years of age |
| 8 | All those 55-59 years of age |
| 9 | All those 50-54 years of age |
| 10a | All those 40-49 years of age |
| 10b | All those 18-39 years of age |

See original priority groups here: https://assets.publishing.service.gov.uk/government/uploads/system/uploads/attachment\_data/file/1007737/Greenbook\_chapter\_14a\_30July2021.pdf#page=15
See revised priority groups here: https://www.england.nhs.uk/coronavirus/wp-content/uploads/sites/52/2021/07/C1327-covid-19-vaccination-autumn-winter-phase-3-planning.pdf

Note that we excluded care home residents and health care workers in
our analysis, so members of JCVI group 1 are not included and JCVI group
2 includes only those aged 80 and over. The original priority group list
has 9 groups, with a 10th group implicitly defined as “everybody else”.
Here we explicitly define this group, and split into two (10a and 10b)
because of the earlier booster eligiblity in the 40-49 group from 15
November 2021 onwards. (https://www.gov.uk/government/news/jcvi-issues-advice-on-covid-19-booster-vaccines-for-those-aged-40-to-49-and-second-doses-for-16-to-17-year-olds)

### Variables and codelists

| Variable | Data source | Notes | Values | Codelist identifier |
| --- | --- | --- | --- | --- |
| Demographics | | | | |
| Sex | Primary care record |  | Male; Female |  |
| Age | Primary care record |  | >=18 |  |
| Ethnicity | Primary care record; SUS-APCS | Taken from primary care record where known, and supplemented by SUS-APCS data where unknown | Black; Mixed; South Asian; White; Other | opensafely/ethnicity/2020-04-27/ |
| NHS region | Primary care record |  | East; London; Midlands; North East and Yorkshire; North West; South East; South West |  |
| English Index of Multiple Deprivation | Primary care record | Derived from Middle Layer Super Output Area (MSOA) of patient's address and using 2019 MSOA rankings | (grouped into 5 categories by IMD rank) |  |
| Care home residency | Primary care record; Care and Quality Commission care home address data | Patients were considered to be care-home residents if they met any one of three criteria used to identify care homes. These criteria are based on either patient address, household age and size, or coded clinical events. See https://wellcomeopenresearch.org/articles/6-90/v1 for more details on how these flags are derived. | 0/1 | primis-covid19-vacc-uptake/longres/v1 primis-covid19-vacc-uptake/carehome/v1 primis-covid19-vacc-uptake/nursehome/v1 primis-covid19-vacc-uptake/domcare/v1 |
| Health and social care worker status | NIMS | Vaccinees were asked if they were a health care worker or carer at the time of vaccination. | 0/1 |  |
|  |  |  | 0/1 |  |
| Clinical | | | | |
| Severe obesity | Primary care record | BMI >= 40, based on most recent weight measurement, or clinically coded severe obesity | 0/1 | primis-covid19-vacc-uptake/sev\_obesity/v1.2 primis-covid19-vacc-uptake/bmi\_stage/v1.2 primis-covid19-vacc-uptake/bmi/v1 |
| Asthma | Primary care record | Based on PRIMIS specification | 0/1 | primis-covid19-vacc-uptake/ast/v1 primis-covid19-vacc-uptake/astadm/v1 primis-covid19-vacc-uptake/astrx/v1 |
| Chronic respiratory disease | Primary care record | Based on PRIMIS specification | 0/1 | primis-covid19-vacc-uptake/resp\_cov/v1 |
| Chronic heart disease | Primary care record | Based on PRIMIS specification | 0/1 | primis-covid19-vacc-uptake/chd\_cov/v1.2.1 |
| Chronic kidney disease | Primary care record | Based on PRIMIS specification | 0/1 | primis-covid19-vacc-uptake/ckd\_cov/v1.2.1 primis-covid19-vacc-uptake/ckd15/v1 primis-covid19-vacc-uptake/ckd35/v1 |
| Chronic liver disease | Primary care record | Based on PRIMIS specification | 0/1 | primis-covid19-vacc-uptake/cld/v1 |
| Diabetes | Primary care record | Based on PRIMIS specification | 0/1 | primis-covid19-vacc-uptake/dmres/v1 primis-covid19-vacc-uptake/diab/v1 |
| Chronic neurological disease | Primary care record | Based on PRIMIS specification | 0/1 | primis-covid19-vacc-uptake/cns\_cov/v1 |
| Immunosuppressed | Primary care record | Based on PRIMIS specification | 0/1 | primis-covid19-vacc-uptake/immdx\_cov/v1 primis-covid19-vacc-uptake/immdx/v1 |
| Asplenia | Primary care record | Based on PRIMIS specification | 0/1 | primis-covid19-vacc-uptake/spln\_cov/v1/ |
| Severe mental illness | Primary care record | Based on PRIMIS specification | 0/1 | primis-covid19-vacc-uptake/sev\_mental/v1 primis-covid19-vacc-uptake/smhres/v1 |
| Learning disabilities | Primary care record | Based on PRIMIS specification | 0/1 | primis-covid19-vacc-uptake/learndis/v1 |
| End-of-life care | Primary care record | Based on codes indicating participation in end-of-life or palliative care pathways, or Midalozam prescriptions | 0/1 | nhsd-primary-care-domain-refsets/palcare\_cod/5fce98cf opensafely/midazolam-end-of-life/4c1b3c89 |
| Housebound | Primary care record |  | 0/1 | opensafely/housebound/5bc77310 primis-covid19-vacc-uptake/carehome/v1 opensafely/no-longer-housebound/29a88ca6 |
| Clinically extremely vulnerable | Primary care record | Based on PRIMIS specification | 0/1 | primis-covid19-vacc-uptake/shield/v1 primis-covid19-vacc-uptake/nonshield/v1 |
| Clinically at-risk | Primary care record | immunosuppressed; asplenia; chronic heart disease; chronic kidney disease; chronic liver diease; diabetes; chronic respiratory disease; asthma; chronic neurological disease; learning disabilities; severe mental illness | 0/1 |  |
| Comorbidity count | Primary care record | Count of: severe obesity; chronic heart disease; chronic kidney disease; chronic liver diease; diabetes; chronic respiratory disease OR asthma; chronic neurological disease | >=0 |  |
| In-hospital status, planned | SUS-APCS | With admission method in ["11", "12", "13", "81"] | Date |  |
| In-hospital status, unplanned | SUS-APCS | With admission method in ["21", "22", "23", "24", "25", "2A", "2B", "2C", "2D", "28"] | Date |  |
| Vaccination | | | | |
| Vaccination status | NIMS / Primary care record | Vaccination details are recorded in the National Immunisation Management Service (NIMS) and transmitted to the patient's GP record within days. Vaccination status can be identified from by the vaccine product name. Vaccine cardinality (i.e., first dose, second etc) is identified by vaccine dates, rather than explicit clinical coding. | Date + BNT162b2; ChAdOx1-S; mRNA-1273 |  |
| COVID-19 events | | | | |
| Probable COVID-19 | Primary care record | Exact dates of COVID-19 infection / disease in priamry care records are unreliable. This variable is only used to determine historical infection prior to the study start date. Important due to low availability of testing early in the pandemic |  | opensafely/covid-identification-in-primary-care-probable-covid-positive-test/2020-07-16/ opensafely/covid-identification-in-primary-care-probable-covid-clinical-code/2020-07-16/ opensafely/covid-identification-in-primary-care-probable-covid-sequelae/2020-07-16/ |
| SARS-CoV-2 positive test | SGSS | Any positive SARS-CoV-2 test, whether PCR or LFT. Swab date is used as the event date, not the date that the result was recorded. | Date |  |
| COVID-19 Hospital admissions | SUS-APCS | Any (completed) hospital episode with COVID-19 ICD10 codes mentioned anywhere in the diagnosis field (not just the primary diagnosis), and with admission method in ["21", "22", "23", "24", "25", "2A", "2B", "2C", "2D", "28"] | Date | opensafely/covid-identification/2020-06-03/ |
| COVID-19 death | Death register (ONS) | Deaths with COVID-19 ICD10 diagnosis codes mentioned anywhere on the death certificate (not just underlying cause) | Date | opensafely/covid-identification/2020-06-03/ |
| NA | | | | |
| Death | Death register (ONS) | Any death | Date |  |

Codelists can be found at
`https://codelists.opensafely.org/codelist/<ID>`,
substituting `<ID>` for the codelist identifier in the
table above.

###### Table S1: Characteristics of candidate participants

Candidate matching characteristics as on the day of recruitment into
the treatment or control group.

| Characteristic | Boosted, eligible, matched | Control |
| --- | --- | --- |
| Total N | 3,426,960 | 3,426,960 |
| Primary vaccine course |  |  |
| BNT162b2-BNT162b2 | 1,476,131 (43%) | 1,476,131 (43%) |
| ChAdOx1-ChAdOx1 | 1,950,829 (57%) | 1,950,829 (57%) |
| Age |  |  |
| 18-39 | 184,832 (5.4%) | 199,354 (5.8%) |
| 40-49 | 221,668 (6.5%) | 242,618 (7.1%) |
| 50-59 | 628,035 (18%) | 632,212 (18%) |
| 60-69 | 939,494 (27%) | 932,775 (27%) |
| 70-79 | 1,039,938 (30%) | 1,011,653 (30%) |
| 80-89 | 364,261 (11%) | 351,385 (10%) |
| 90+ | 48,732 (1.4%) | 56,963 (1.7%) |
| Sex |  |  |
| Female | 1,860,622 (54%) | 1,865,153 (54%) |
| Male | 1,566,338 (46%) | 1,561,807 (46%) |
| Ethnicity |  |  |
| White | 3,218,953 (94%) | 3,195,436 (93%) |
| Black | 31,058 (0.9%) | 37,879 (1.1%) |
| South Asian | 126,998 (3.7%) | 145,055 (4.2%) |
| Mixed | 18,073 (0.5%) | 19,164 (0.6%) |
| Other | 31,878 (0.9%) | 29,426 (0.9%) |
| IMD |  |  |
| 1 most deprived | 469,397 (14%) | 558,365 (16%) |
| 2 | 586,546 (17%) | 641,493 (19%) |
| 3 | 763,813 (22%) | 775,775 (23%) |
| 4 | 799,431 (23%) | 750,914 (22%) |
| 5 least deprived | 807,773 (24%) | 700,413 (20%) |
| Region |  |  |
| North East and Yorkshire | 616,628 (18%) | 616,628 (18%) |
| Midlands | 774,777 (23%) | 774,777 (23%) |
| North West | 285,853 (8.3%) | 285,853 (8.3%) |
| East of England | 825,941 (24%) | 825,941 (24%) |
| London | 109,498 (3.2%) | 109,498 (3.2%) |
| South East | 261,876 (7.6%) | 261,876 (7.6%) |
| South West | 552,387 (16%) | 552,387 (16%) |
| Clinically extremely vulnerable | 470,981 (14%) | 470,981 (14%) |
| Body Mass Index > 40 kg/m^2 | 156,646 (4.6%) | 164,724 (4.8%) |
| Chronic heart disease | 665,488 (19%) | 659,164 (19%) |
| Chronic kidney disease | 316,348 (9.2%) | 315,120 (9.2%) |
| Diabetes | 532,340 (16%) | 562,011 (16%) |
| Chronic liver disease | 115,145 (3.4%) | 121,472 (3.5%) |
| Chronic respiratory disease | 249,441 (7.3%) | 252,359 (7.4%) |
| Asthma | 25,437 (0.7%) | 24,184 (0.7%) |
| Chronic neurological disease | 274,335 (8.0%) | 292,425 (8.5%) |
| Immunosuppressed | 154,260 (4.5%) | 121,969 (3.6%) |
| Asplenia or poor spleen function | 40,134 (1.2%) | 36,303 (1.1%) |
| Learning disabilities | 21,039 (0.6%) | 26,533 (0.8%) |
| Serious mental illness | 31,996 (0.9%) | 42,422 (1.2%) |
| Number of SARS-CoV-2 tests |  |  |
| 0 | 2,092,836 (61%) | 2,217,161 (65%) |
| 1 | 486,362 (14%) | 467,274 (14%) |
| 2 | 206,142 (6.0%) | 193,982 (5.7%) |
| 3+ | 641,620 (19%) | 548,543 (16%) |
| Prior documented SARS-CoV-2 infection | 202,432 (5.9%) | 202,432 (5.9%) |
| In hospital (planned admission) | 46,478 (1.4%) | 46,421 (1.4%) |

###### Figure S1: Booster uptake and matching success

Cumulative booster vaccination over the duration of the study period,
by matching eligibility and matching success.

##### Figure S2: Standardised Mean Differences between Boosted and Unboosted groups

For variables that were matched exactly the SMD is zero, and so are
not shown.

#### Figure S3: Estimated booster effectiveness, all models
